# Ivabradine Treatment to Prevent Anthracycline-Induced Cardiotoxicity: A Randomized Clinical Trial

**DOI:** 10.1101/2024.10.30.24316463

**Authors:** Stephanie Itala Rizk, Isabela Bispo Santos da Silva Costa, Cecilia Beatriz Bittencourt Viana Cruz, Brunna Pileggi, Fernanda Thereza de Almeida Andrade, Thalita Barbosa Gonzalez, Cristina Salvadori Bittar, Julia Tizue Fukushima, Vinicius Caldeira Quintao, Eduardo Atsushi Osawa, Juliana Barbosa Sobral Alves, Silvia Moulin Ribeiro Fonseca, Diego Ribeiro Garcia, Juliana Pereira, Valeria Buccheri, Juliana Ávila, Lucas Tokio Kawahara, Cecilia Chie Sakaguchi Barros, Lucas Takeshi Ikeoka, Letícia Naomi Nakada, Mariella Fellini, Vanderson Geraldo Rocha, Eduardo Magalhães Rego, Paulo Marcelo Gehm Hoff, Roberto Kalil Filho, Giovanni Landoni, Ludhmila Abrahão Hajjar

## Abstract

**Background:** Cancer therapy-related cardiac dysfunction frequently occurs in patients receiving anthracycline. Ivabradine reduces the heart rate without affecting contractility and has shown anti-inflammatory, antioxidant, and antiapoptotic effects in experimental models of cardiotoxicity. This study aims to evaluate the effect of ivabradine on cancer therapy-related cardiac dysfunction incidence in patients with lymphoma or sarcoma treated with anthracycline.

**Methods:** This study was a prospective, randomized, and triple-blind trial. Patients starting anthracycline therapy were given either ivabradine 5 mg twice daily or placebo until 30 days after completing treatment. The primary outcome was a ≥10% relative reduction in global longitudinal strain at 12 months. The secondary outcomes included 12-month clinical outcomes, a ≥10% decrease in the left ventricular ejection fraction to <55%, diastolic dysfunction, and troponin T and NT-proBNP levels.

**Results:** This study was conducted with 107 patients (51 in the ivabradine group and 56 in the placebo group). The median dose of anthracycline was 300 mg/m^2^ (250--300 mg/m^2^) in both groups. A ≥10% relative reduction in global longitudinal strain at 12 months was reached in 57% of the ivabradine group and in 50% of the placebo group (OR 1.32, 95% CI: -0.612.83, p=.477). Fewer patients in the ivabradine group than in the placebo group had troponin T levels ≥14 ng/L [16 (39.0%) vs. 23 (62.2%), p=0.041] at 6 months. There were no differences in the other secondary outcomes.

**Conclusions:** A fixed 10 mg/day dose of ivabradine does not protect cancer patients against anthracycline cardiotoxicity.

**Clinical trials registration:** NCT03650205 https://clinicaltrials.gov/study/NCT03650205?cond=NCT03650205&rank=1

**Clinical perspectives:**

- This study found that ivabradine at a fixed dose of 10 mg/day does not effectively prevent cardiotoxicity in cancer patients undergoing anthracycline (ANT) therapy, contrary to previous expectations.
- These findings suggest that ivabradine may not be sufficient as a cardioprotective agent in ANT therapy, emphasizing the need to explore additional or alternative strategies to manage cardiotoxicity in cancer patients.
- The study highlights a potentially complex interaction between ANT and cardiac tissue, indicating a need for further research to fully understand and address this risk.

## Introduction

While effective at targeting and destroying cancer cells, chemotherapy can also have unintended detrimental effects on the cardiovascular system.^1^ These include an increased risk of heart failure (HF), hypertension, arrhythmias, myocardial ischemia, and thromboembolism. These issues are often referred to as cancer therapy-related cardiovascular dysfunction (CTRCD) or cardiotoxicity.^2^

Anthracyclines are highly effective in treating various cancers, including breast cancer and hematologic malignancies, but are highly likely to compromise the cardiovascular system. The risk of CTRCD from anthracycline (ANT) is dose-related, with higher cumulative doses increasing the risk of cardiac damage, which primarily causes myocardial injury and HF.^3^ However, even low doses of anthracycline can still be risky, particularly for vulnerable groups such as elderly individuals or those with preexisting heart disease. ANT-induced cardiotoxicity is complex and involves oxidative stress, DNA damage, and disruption of cardiomyocyte metabolism.^4^

The management of CTRCD in patients receiving ANT therapy is challenging. Randomized studies have not shown consistent effects of beta-blockers, angiotensin-converting enzyme inhibitors and angiotensin receptor blockers in the prevention of ANT cardiotoxicity.^5^

Experimental studies investigating the use of ivabradine for the prevention of ANT-associated cardiotoxicity have demonstrated anti-inflammatory and antioxidant effects and reduced plasma levels of tumor necrosis factor alpha, troponin, lactate dehydrogenase, and malondialdehyde.^6,7^ Furthermore, an elevated heart rate (HR) is a recognized marker of cardiovascular risk and is linked to atherosclerosis progression and cardiac dysfunction due to imbalances in oxygen supply and demand, endothelial shear stress, and enzyme activation.^8–10^ Colin et al demonstrated that a reduction in HR reduces oxygen consumption through decreased stress on the left ventricular (LV) wall and increased diastolic time.^11^ Ivabradine selectively modulates the HR, without affecting cardiac inotropy, specifically by inhibiting the I(f) current in the sinoatrial node.

This Ivabradine to Prevent Anthracycline-Related Cardiovascular Dysfunction (IPAC) trial was a randomized clinical trial conducted to assess the effectiveness of ivabradine in preventing cardiovascular dysfunction in lymphoma or sarcoma patients treated with ANT.

## Methods

### Study population

The *IPAC* trial was a triple-blind, randomized, placebo-controlled study conducted at the Instituto do Câncer do Estado de São Paulo (ICESP) and at the Instituto do Coração (InCor) do Hospital das Clínicas da Faculdade de Medicina da Universidade de São Paulo, Brazil, after approval by the Research Ethics Committee. Consecutive patients with lymphoma or sarcoma slated for ANT chemotherapy were assessed for eligibility. The participants were informed, provided consent, signed up for the study, and were registered on *ClinicalTrials.gov* (NCT03650205). Funding was provided by the Sao Paulo Research Foundation.

The exclusion criteria were an inability to assess LV function, prior chemotherapy with anthracycline or radiation, HF symptoms, existing cardiomyopathy, coronary or valve disease, atrial fibrillation, bradycardia, chronic renal disease, a positive test result indicating SARS-CoV-2 infection and allergy/contraindication to ivabradine.

### Study design

Eligible patients were enrolled and randomly assigned to receive either ivabradine 5 mg twice daily or placebo, starting with chemotherapy initiation and continuing through the ANT regimen until 30 days after therapy. Randomization was performed via a computer system, with data held by an independent research pharmacy. Ivabradine was carefully encapsulated so that it was visually indistinguishable from the placebo. The participants, healthcare professionals, data managers and statisticians were blinded to the treatment assignments.

### Study procedures

The enrolled patients underwent medical evaluation; laboratory assessments, including cardiac biomarkers [cardiac troponin T and NT-proBNP]; electrocardiogram (EKG); transthoracic echocardiogram (TTE), featuring myocardial global longitudinal strain (GLS) measurements; and 24-hour *Holter* monitoring at baseline (prior to the initiation of chemotherapy). For patients who met the eligibility criteria, randomization was performed, and the medication (ivabradine or placebo) was initiated on the first day of chemotherapy.

A standard global tool for assessing health and quality of life and analyzing mobility, self-care, usual activities, pain/discomfort, and anxiety/depression (EQ-5D-3L questionnaire) was administered at baseline and after chemotherapy.

Sequential laboratory testing, EKG assessments, and TTE examinations of the strains were conducted at 3, 6 and 12 months following the initiation of chemotherapy. *Holter* monitoring was also performed at the conclusion of the treatment. An overview of the study procedures is provided in Figure 1. TTE was conducted employing a commercially available system with digital ultrasonic equipment (Vivid 9, GE Healthcare, Milwaukee, USA), and all measurements adhered to the recommendations provided by the American Society of Echocardiography.^12^

**Figure 1.**
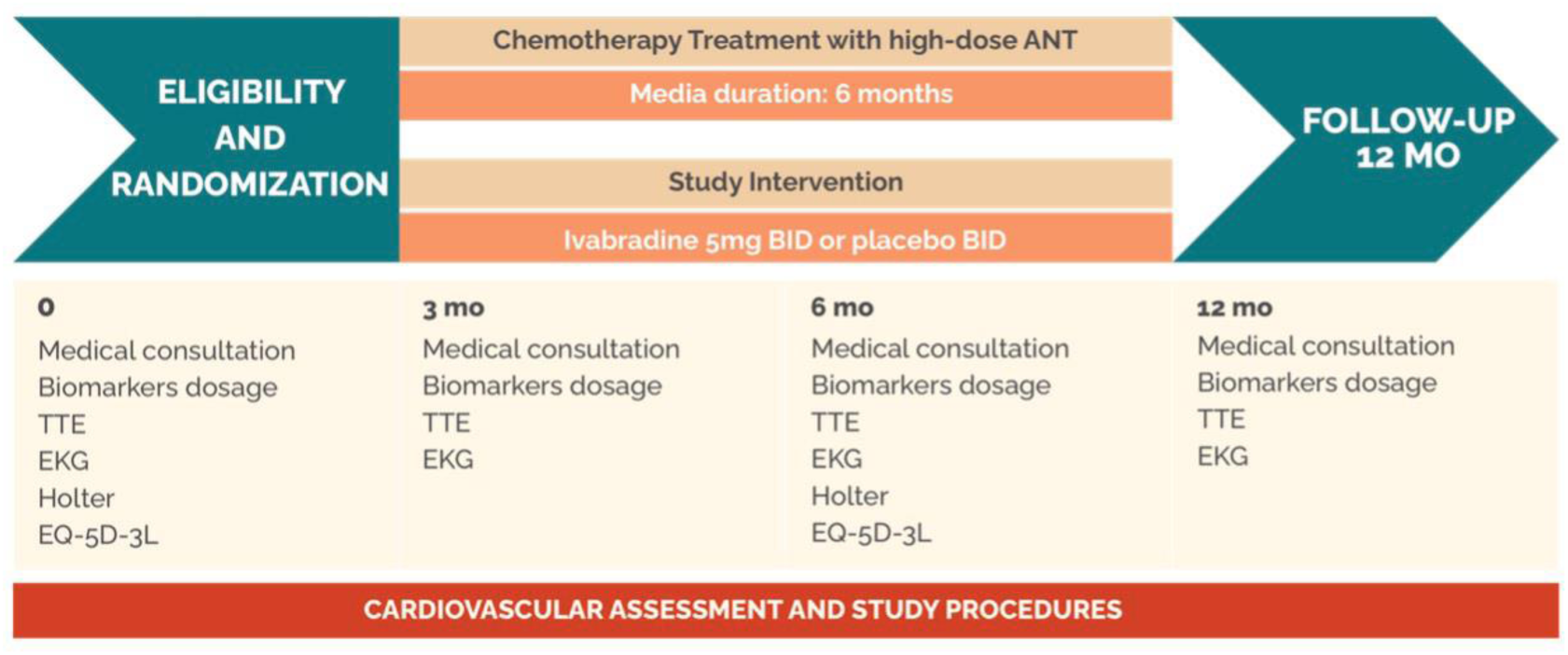
**Title:** Study procedures and assessment timeline **Caption:** The timeline and key medical assessments conducted at baseline (0 months), during the chemotherapy treatment phase with high-dose ANT (3 and 6 months), and during the 12-month follow-up after intervention are summarized. The schedule included medical consultations, biomarker dosages, transthoracic echocardiogram, electrocardiogram, Holter monitoring, and EQ-5D-3L assessments at each designated time point (0, 3, 6, and 12 months). ANT: anthracycline, BID: *bis in die,* MO: months, TTE: transthoracic echocardiographic, EKG: electrocardiogram, EQ-5D-3L: EuroQol-5 Dimension-3 Levels.

The left ventricular ejection fraction (LVEF) was measured by Simpson’s method via apical 4- and 2-chamber views. Myocardial GLS analysis was performed via semiautomated speckle tracking, covering the whole LV from three apical views for the assessment of cardiac cycle tissue deformation. The endocardial border of the LV was manually traced at end-systole and autoadjusted to include the full myocardium.^13,14^

The diastolic function evaluation included an assessment of the mitral inflow E/A pattern, E/A ratio, E velocity deceleration time, annular tissue *Doppler* curves (é/á), and E/é ratio. Additional echocardiographic parameters that were assessed via Doppler echocardiography included the left atrium diameter and volume, interventricular septum diameter, posterior wall thickness, LV end-diastolic diameter, LV end-systolic diameter, and mitral inflow.^15^ Two experienced and board-certified echocardiographers performed the exams. Diagnosing LV diastolic dysfunction involves evaluating the E/e’ ratio, e’ wave velocity, indexed left atrial volume and tricuspid regurgitation velocity. Normal diastolic function is defined by alterations in fewer than 50% of these criteria; diastolic dysfunction, by alterations in more than 50% of the criteria; and indeterminate diastolic function, by alterations in exactly 50% of the criteria. Grade I dysfunction (normal left atrial pressure) is indicated by a specific E/A ratio and E wave values, and grade III dysfunction (elevated left atrial pressure) is indicated by a higher E/A ratio. Intermediate E/A ratio values require additional criteria assessments. The classification of dysfunction as grade I or II depends on the number of positive or negative criteria, whereas a split evaluation results in an indeterminate classification.^16^

Arrhythmia was diagnosed when any of the following conditions were present: ventricular arrhythmia, ventricular fibrillation, ventricular tachycardia, atrial fibrillation, atrial flutter, atrial tachycardia, supraventricular tachycardia, atrioventricular block, conduction disorder, or sick sinus syndrome.^17^

Ultrasensitive troponin T was measured via an advanced electrochemiluminescence method. In this assay, troponin T antibodies are labeled with ruthenium complexes that emit light when activated by an electrical impulse. The threshold for abnormal troponin T levels was set at ≥14 ng/L, which is consistent with current clinical guidelines, to ensure relevance to the patient population.^18^

The measurement of NT-proBNP levels was carried out via a highly sensitive and specific microparticle immunoassay method that employs chemiluminescence. The chemiluminescent reaction emits light proportionally to the NT-proBNP concentration present. For clinical relevance, a reference value of up to 125 pg/mL was established, which is considered the upper limit of normal for NT-proBNP levels and aids in the interpretation of the assay results in the context of cardiac function and potential heart failure.^19^

### Study outcomes

The primary outcome was a relative reduction of at least 10% in the value of GLS through TTE 12 months after the start of the study.^20,21^

The secondary outcomes were a combined endpoint of all-cause death, acute myocardial infarction, HF, and arrhythmias at 12 months; a reduction in LVEF of at least 10%, resulting in an LVEF below 55% and a change in diastolic dysfunction at 12 months; the quantification of troponin T and NT-proBNP levels at 3, 6, and 12 months; and a change in GLS at 180 days. We also analyzed the adverse effects of treatment at 12 months.

### Statistical analysis

The initial sample size was calculated with an expected incidence of cardiotoxicity of 50% with the use of ANT and an expected reduction to 25% with ivabradine.^22^ With a statistical significance level of 95% and to achieve 90% statistical power with a 2-sided Fisher exact test, 160 patients (80 in each arm) were needed. Owing to recruitment challenges, mainly caused by pandemics, the sample size was revised. We adjusted for 80% power, keeping the same endpoints and hypothesis, and the required number of patients was recalculated to 100 to detect a reduction in the proportion of patients with cardiotoxicity from 50% to 25%.

Descriptive statistics were used for the distribution of variables; continuous variables are summarized herein as the means with standard deviations or as medians with interquartile ranges, and categorical variables are summarized as counts and percentages. All the statistical analyses were performed on intention-to-treat. Continuous variables were compared via t tests or Mann‒Whitney U tests, and categorical variables were compared via the Pearson χ^2^, Fisher exact or likelihood ratio test. Variables measured at multiple time points were evaluated via repeated measures analysis of variance or the Mann‒Whitney U test and Wilcoxon signed-rank test. For the analysis of categorical outcomes, differences between groups and odds ratios were calculated, along with their respective 95% confidence intervals.

The statistical analysis was conducted via SPSS version 25.0 (Statistical Package for the Social Sciences), and p values less than 0.05 were considered significant.

## Results

### Patient characteristics

Between January 2019 and May 2022, a total of 270 patients with a diagnosis of lymphoma or sarcoma were screened. We randomized 107 patients to receive ivabradine (n=51) or placebo (n=56) for the intention-to-treat analysis (Figure 2).

**Figure 2.**
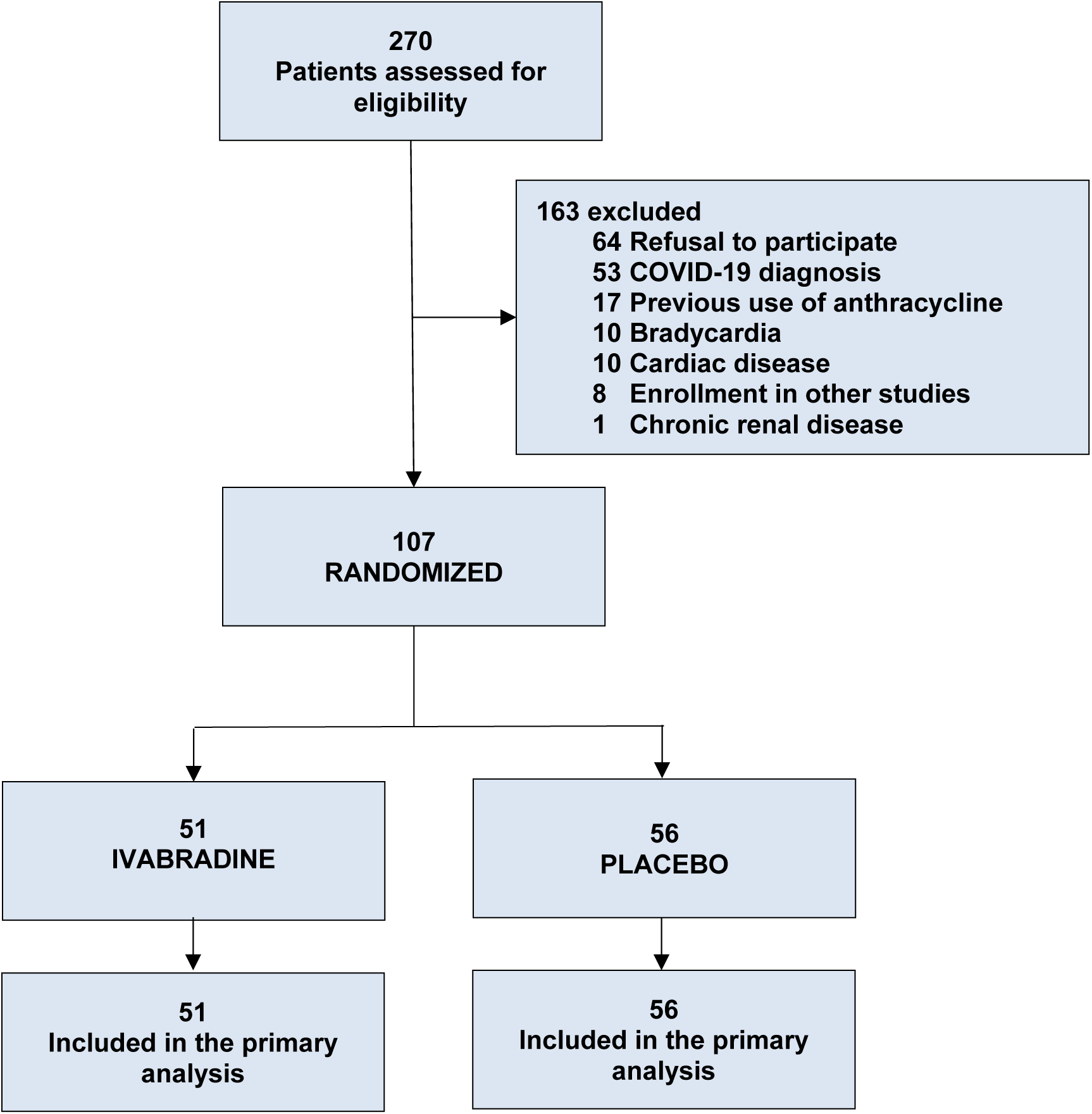
**Title**: Study flowchart. **Caption:** Depiction of the participant allocation from the initial assessment to the completion of analysis within the study arms.

The baseline characteristics were well balanced between the groups (Table 1). Most patients had lymphoma, and the median ANT dose was 300 mg/m^2^ (250–300 mg/m^2^) in both groups.

**Table 1.** Baseline and demographic characteristics of the patients.

| Variable | Ivabradine<br>(n=51) | Placebo<br>(n=56) |
| --- | --- | --- |
| Sex | 18 (35.3%) | 28 (50.0%) |
| Female |  |  |
| Age (years), median (IQR) | 49 (32-59) | 39 (26-60) |
| Race |  |  |
| White | 37 (72.5%) | 45 (80.4%) |
| Black | 3 (5.9%) | 4 (7.1%) |
| Other/unknown | 11 (21.6%) | 7 (12.5%) |
| Smoking |  |  |
| Former smoker | 13 (25.5%) | 6 (10.7%) |
| Current smoker | 7 (13.7%) | 9 (16.1%) |
| Alcohol consumption |  |  |
| Social/sporadic | 17 (33.3%) | 20 (35.7%) |
| Daily | 4 (7.8%) | 3 (5.4%) |
| Sedentary lifestyle | 34 (66.7%) | 35 (62.5%) |
| Diabetes | 4 (7.8%) | 5 (8.9%) |
| Hypertension | 13 (25.5%) | 9 (16.4%) |
| Dyslipidemia | 4 (8.0%) | 3 (5.4%) |
| Diagnosis |  |  |
| Diffuse NHL | 23 (45.1%) | 23 (41.1%) |
| Follicular NHL | 7 (13.7%) | 6 (10.7%) |
| HL | 18 (35.3%) | 26 (46.4%) |
| Sarcoma | 3 (5.9%) | 1 (1.8%) |
| Anthracycline dose*<br>(mg/m<sup>2</sup>), median (IQR) | 300 (250-300) | 300 (250-300) |

a: Pearson's chi-square test, b: likelihood ratio test, c: Fisher's exact test, d: Mann–Whitney test.
\*Doxorubicin.
IQR: Interquartile range; BCC: calcium channel blocker; ARB: angiotensin 2 receptor
blocker; CAD: coronary artery disease; ECOG: Eastern Cooperative Oncology Group; ACE
inhibitors: angiotensin converting enzyme inhibitors; NHL: non-Hodgkin's lymphoma; LH:
Hodgkin's lymphoma.

### Outcomes

#### Primary outcome

A reduction in GLS of at least 10% at the 12-month follow-up was observed in 29 patients (57%) in the ivabradine group and in 28 patients (50%) in the placebo group [(odds ratio: 1.32; 95% CI: 0.61-2.83), p=0.47] (Table 2). At 3, 6 and 12 months of follow-up, there was a significant reduction in GLS compared with baseline in both groups (p=0.034, p<0.001 and p<0.001), but there was no significant difference between the ivabradine and placebo groups (Figure 3).

**Table 2.**
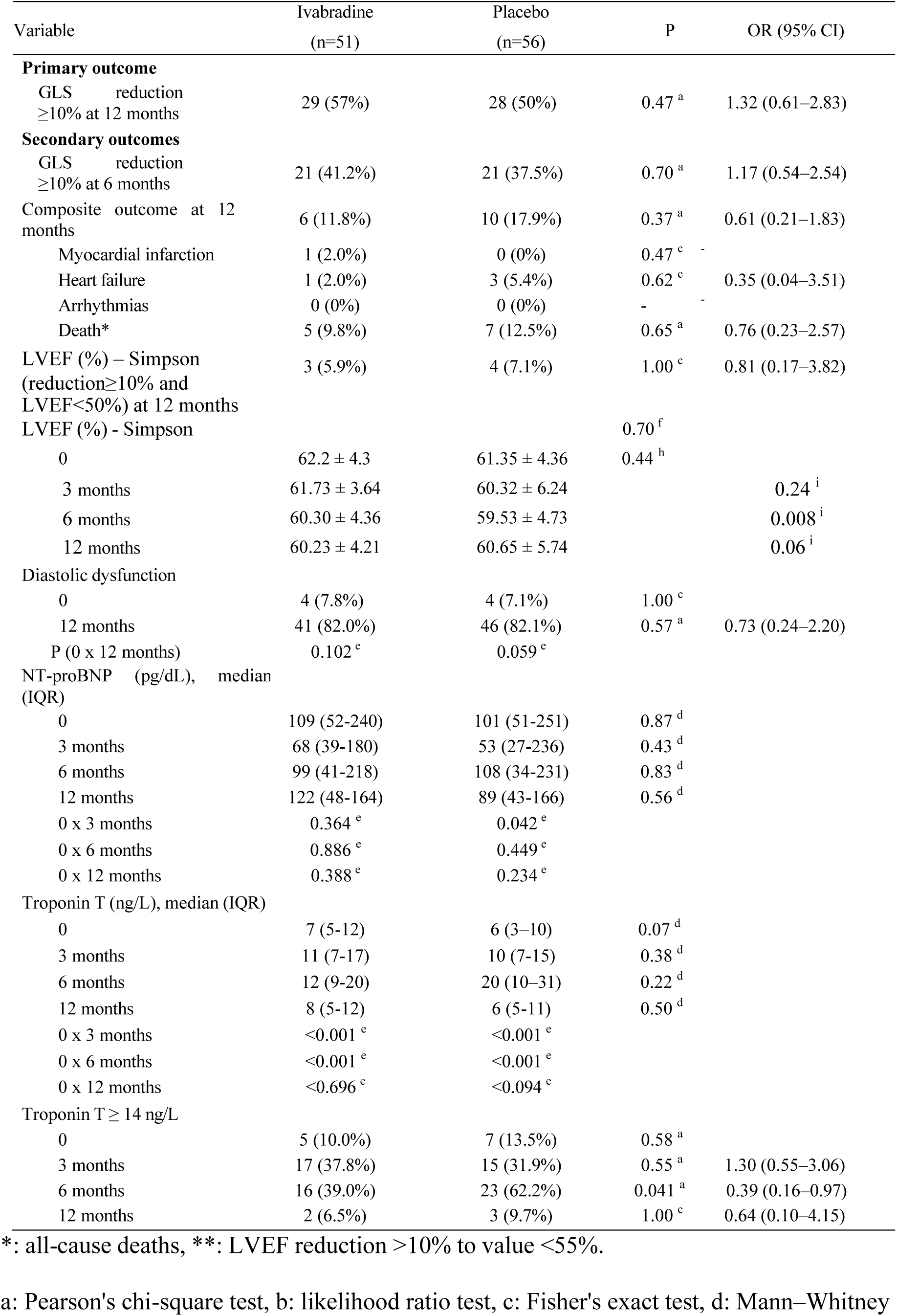
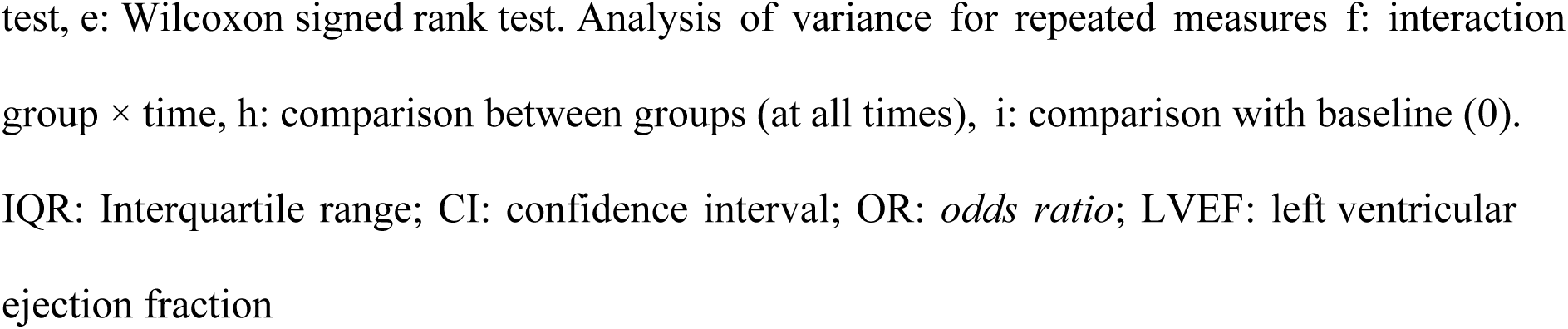
Outcomes of the study.

| Variable | Ivabradine<br>(n=51) | Placebo<br>(n=56) | P | OR (95% CI) |
| --- | --- | --- | --- | --- |
| **Primary outcome** |  |  |  |  |
| GLS reduction<br>≥10% at 12 months | 29 (57%) | 28 (50%) | 0.47<sup>a</sup> | 1.32 (0.61–2.83) |
| **Secondary outcomes** |  |  |  |  |
| GLS reduction<br>≥10% at 6 months | 21 (41.2%) | 21 (37.5%) | 0.70<sup>a</sup> | 1.17 (0.54–2.54) |
| Composite outcome at 12<br>months | 6 (11.8%) | 10 (17.9%) | 0.37<sup>a</sup> | 0.61 (0.21–1.83) |
| Myocardial infarction | 1 (2.0%) | 0 (0%) | 0.47<sup>c</sup> | - |
| Heart failure | 1 (2.0%) | 3 (5.4%) | 0.62<sup>c</sup> | 0.35 (0.04–3.51) |
| Arrhythmias | 0 (0%) | 0 (0%) | - | - |
| Death* | 5 (9.8%) | 7 (12.5%) | 0.65<sup>a</sup> | 0.76 (0.23–2.57) |
| LVEF (%) – Simpson<br>(reduction ≥10% and<br>LVEF <50%) at 12 months | 3 (5.9%) | 4 (7.1%) | 1.00<sup>c</sup> | 0.81 (0.17–3.82) |
| LVEF (%) - Simpson |  |  | 0.70<sup>f</sup> |  |
| 0 | 62.2 ± 4.3 | 61.35 ± 4.36 | 0.44<sup>h</sup> |  |
| 3 months | 61.73 ± 3.64 | 60.32 ± 6.24 |  | 0.24<sup>i</sup> |
| 6 months | 60.30 ± 4.36 | 59.53 ± 4.73 |  | 0.008<sup>i</sup> |
| 12 months | 60.23 ± 4.21 | 60.65 ± 5.74 |  | 0.06<sup>i</sup> |
| Diastolic dysfunction |  |  |  |  |
| 0 | 4 (7.8%) | 4 (7.1%) | 1.00<sup>c</sup> |  |
| 12 months | 41 (82.0%) | 46 (82.1%) | 0.57<sup>a</sup> | 0.73 (0.24–2.20) |
| P (0 x 12 months) | 0.102<sup>e</sup> | 0.059<sup>e</sup> |  |  |
| NT-proBNP (pg/dL), median<br>(IQR) |  |  |  |  |
| 0 | 109 (52-240) | 101 (51-251) | 0.87<sup>d</sup> |  |
| 3 months | 68 (39-180) | 53 (27-236) | 0.43<sup>d</sup> |  |
| 6 months | 99 (41-218) | 108 (34-231) | 0.83<sup>d</sup> |  |
| 12 months | 122 (48-164) | 89 (43-166) | 0.56<sup>d</sup> |  |
| 0 x 3 months | 0.364<sup>e</sup> | 0.042<sup>e</sup> |  |  |
| 0 x 6 months | 0.886<sup>e</sup> | 0.449<sup>e</sup> |  |  |
| 0 x 12 months | 0.388<sup>e</sup> | 0.234<sup>e</sup> |  |  |
| Troponin T (ng/L), median (IQR) |  |  |  |  |
| 0 | 7 (5-12) | 6 (3-10) | 0.07<sup>d</sup> |  |
| 3 months | 11 (7-17) | 10 (7-15) | 0.38<sup>d</sup> |  |
| 6 months | 12 (9-20) | 20 (10-31) | 0.22<sup>d</sup> |  |
| 12 months | 8 (5-12) | 6 (5-11) | 0.50<sup>d</sup> |  |
| 0 x 3 months | <0.001<sup>e</sup> | <0.001<sup>e</sup> |  |  |
| 0 x 6 months | <0.001<sup>e</sup> | <0.001<sup>e</sup> |  |  |
| 0 x 12 months | <0.696<sup>e</sup> | <0.094<sup>e</sup> |  |  |
| Troponin T ≥ 14 ng/L |  |  |  |  |
| 0 | 5 (10.0%) | 7 (13.5%) | 0.58<sup>a</sup> |  |
| 3 months | 17 (37.8%) | 15 (31.9%) | 0.55<sup>a</sup> | 1.30 (0.55–3.06) |
| 6 months | 16 (39.0%) | 23 (62.2%) | 0.041<sup>a</sup> | 0.39 (0.16–0.97) |
| 12 months | 2 (6.5%) | 3 (9.7%) | 1.00<sup>c</sup> | 0.64 (0.10–4.15) |

\*: all-cause deaths, \*\*: LVEF reduction >10% to value <55%.
a: Pearson's chi-square test, b: likelihood ratio test, c: Fisher's exact test, d: Mann–Whitney
test, e: Wilcoxon signed rank test. Analysis of variance for repeated measures f: interaction group × time, h: comparison between groups (at all times), i: comparison with baseline (0). IQR: Interquartile range; CI: confidence interval; OR: *odds ratio*; LVEF: left ventricular ejection fraction

**Figure 3.**
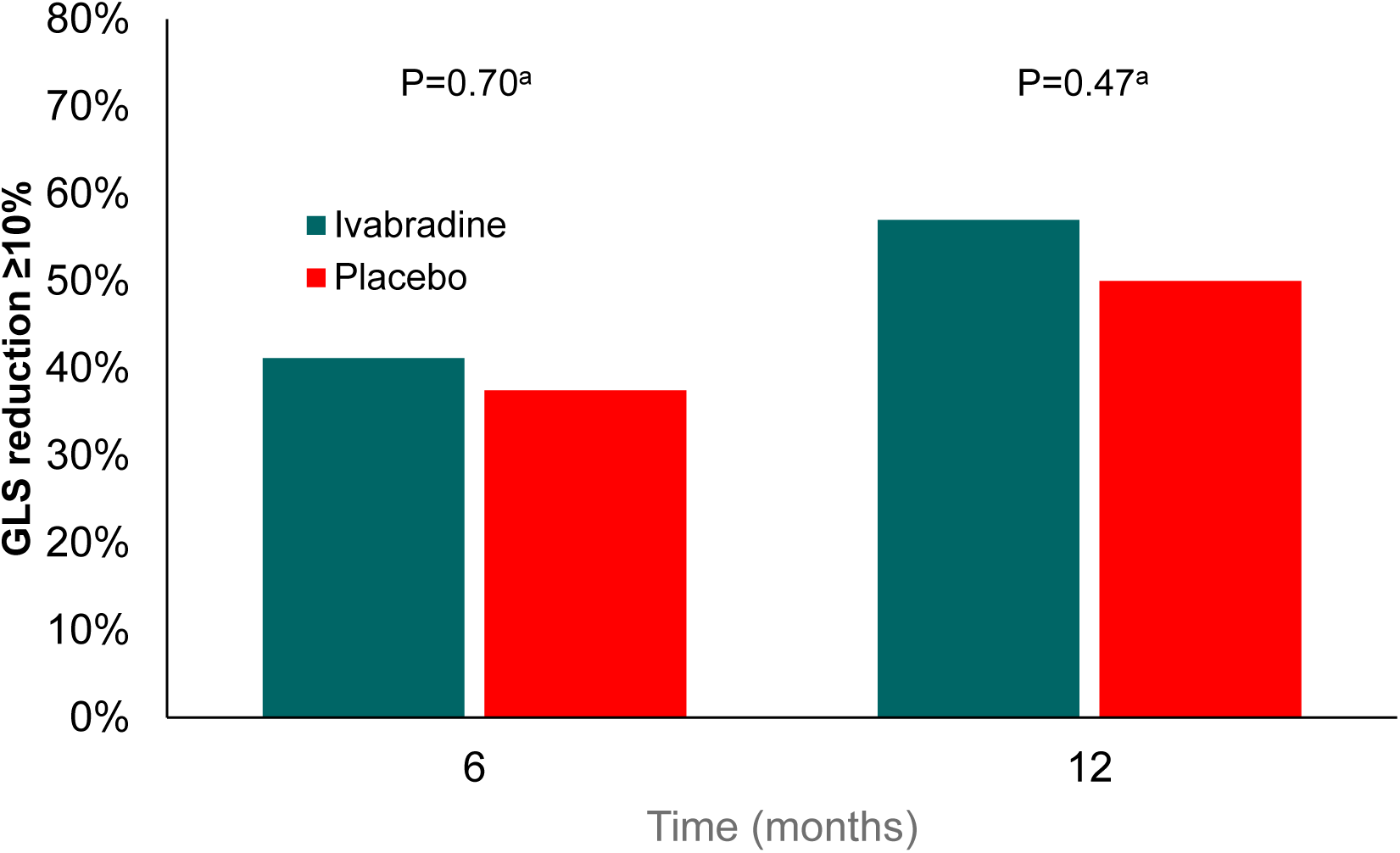
**Title**: Global longitudinal strain of the left ventricle during the study. **Caption:** Global longitudinal strain (GLS) GLS at 3, 6 and 12 months after randomization in comparison with baseline.

#### Secondary outcomes

No significant difference in the occurrence of clinical complications at 12 months was detected between the ivabradine and placebo groups [11.8% vs. 17.9%, OR: 0.61 (95% CI: 0.21–1.83), P=0.37] (Table 2).

Compared with the placebo group, the ivabradine group had 5 deaths (9.8%), whereas the placebo group had 7 deaths (12.5%) [OR: 0.76 (95% CI: 0.23-2.57), P=0.65]. When temporal variations in NT-proBNP levels were considered, there were no statistically significant differences between the studied groups over the 12 months of follow-up (Table 2 and Figure 4A). The comparison of baseline levels with those at 3 months post-intervention revealed a significant reduction only in the placebo group (P=0.042), whereas in the ivabradine group, the change was not statistically significant (P=0.36) (Table 2 and Figure 4A).

**Figure 4.**
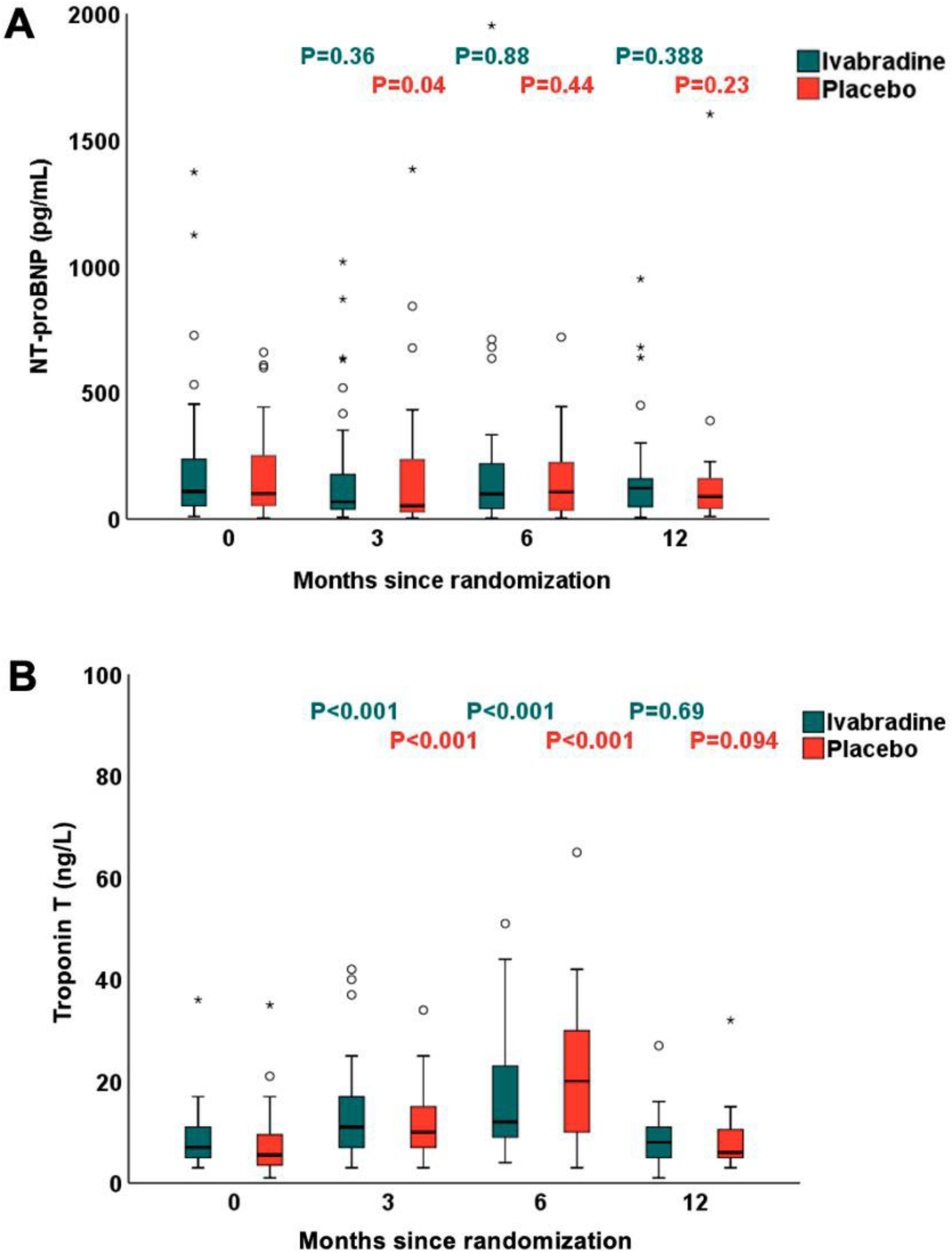
**Title**: Cardiac biomarkers during the study. **Caption:** (A) NT-proBNP (B) Troponin T. P: Wilcoxon signed-rank test (comparison between groups at the baseline value). NT-proBNP: N-terminal pro-B-type natriuretic peptide.

In the ivabradine and placebo groups, the median high-sensitivity troponin T levels at 3 months and 6 months were greater than those at baseline. However, there was no difference between the groups in the troponin T values at any time point (Table 2 and Figure 4B). At 6 months of evaluation, a smaller proportion of patients in the ivabradine group had troponin T levels ≥ 14 ng/L [(16 (39.0%) vs. 23 (62.2%), p=0.041] (Table 2).

The number of patients with a > 10% decrease in LVEF to less than 55% was 3 (5.9%) in the ivabradine group and 4 (7.1%) in the placebo group [odds ratio: 0.81; (95% CI: 0.17-3.82, p=1.00)] (Table 2). The incidence of diastolic dysfunction was greater at 12 months than at baseline in the ivabradine and placebo groups, but there was no significant difference between the groups (17.5% vs. 7.8% in the ivabradine group and 22.5% vs. 7.1% in the placebo group, p=0.73) (Table 2).

There was no significant difference between the groups regarding the HR at 3, 6 and 12 months (p=0.21). However, a reduction in HR was observed in both the ivabradine and placebo groups at 3, 6 and 12 months compared with baseline (Table 3). Ivabradine use was not associated with an increased rate of adverse events. Phosphenes were diagnosed in 3.9% of the ivabradine patients and 1.8% of the placebo patients, whereas bradycardia was diagnosed in 3.9% of the ivabradine patients but in none of the patients receiving placebo (P=0.17). Figure 5 shows the main outcomes of ivabradine treatment in ANT-treated patients.

**Table 3.**
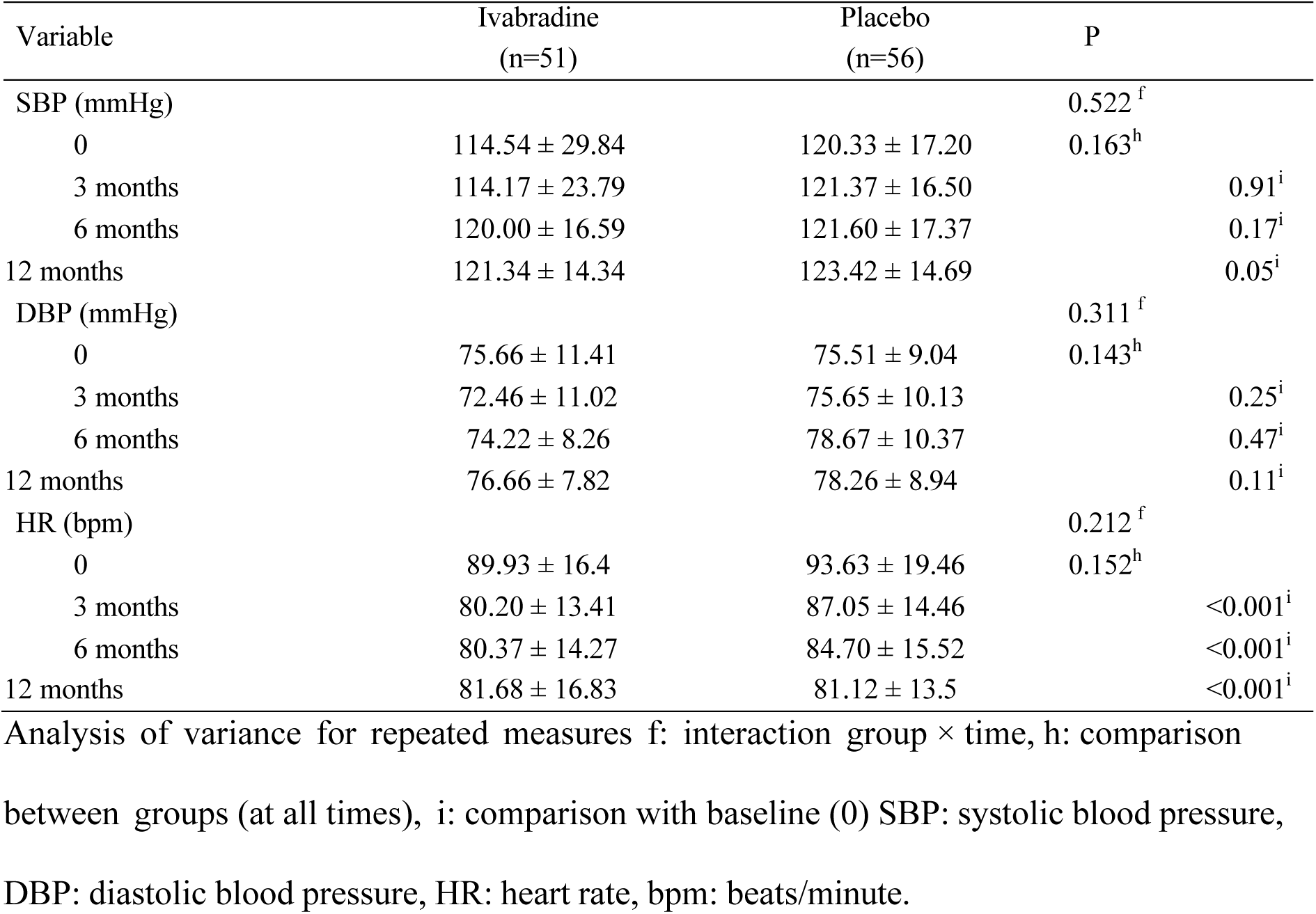
Hemodynamic parameters of the patients.

**Figure 5.**
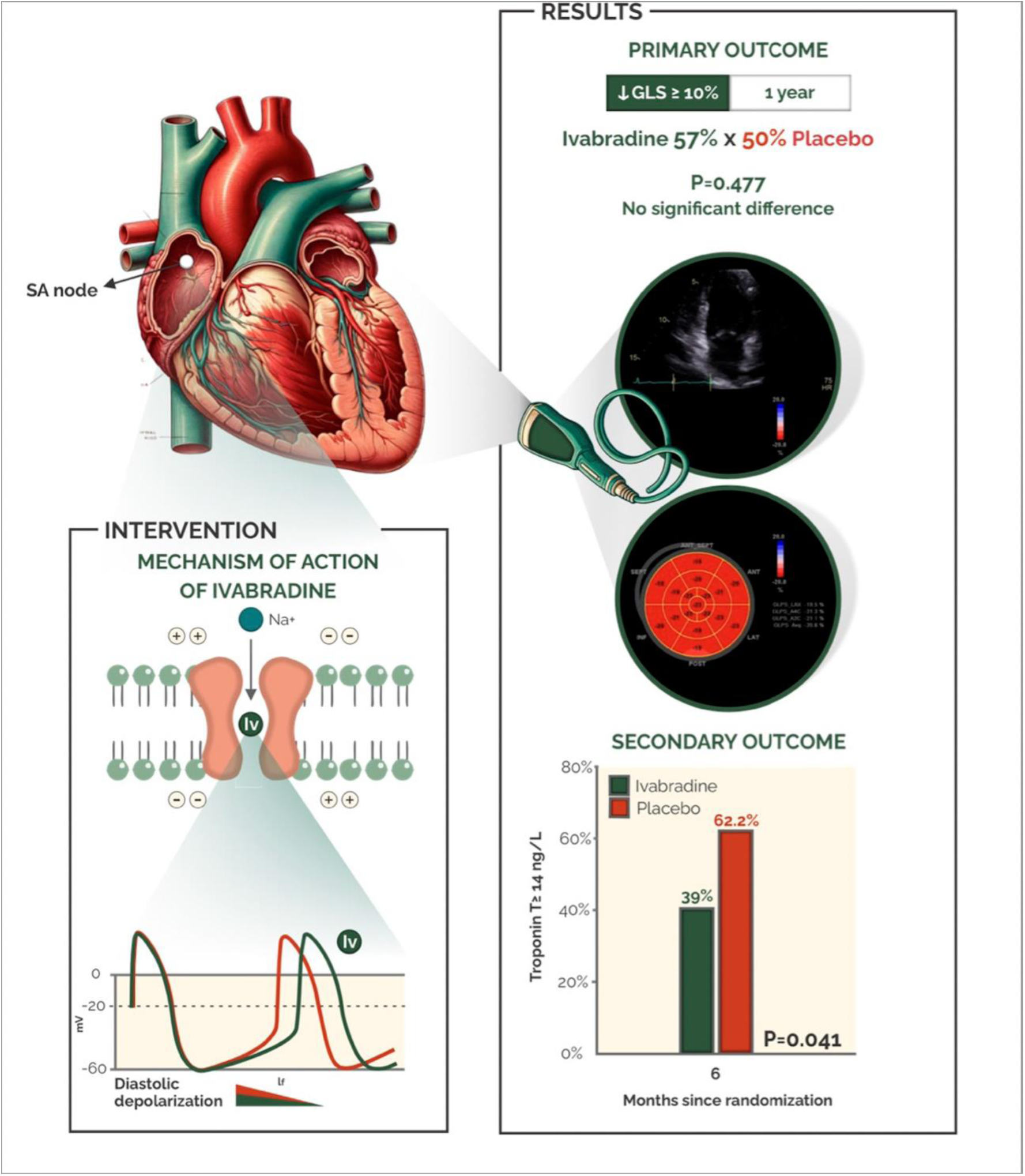
**Title**: Ivabradine treatment to prevent anthracycline-induced cardiotoxicity. **Caption**: The diagram provides a schematic representation of the mechanism of action of ivabradine at the cellular level. Ivabradine treatment in patients with lymphoma or sarcoma treated with anthracycline did not prevent the decline in global longitudinal strain at 12 months.

## Discussion

In patients with lymphoma or sarcoma receiving high-dose anthracycline, ivabradine at a fixed dosage of 10 mg/day did not prevent CTRCD. This is the first published trial designed to evaluate the potential of ivabradine for the prevention of cardiotoxicity in patients receiving ANT. We also observed that at 6 months of follow-up, the number of patients with elevated troponin T levels was lower in the ivabradine group than in the placebo group. Similarly, the CECCY study revealed that carvedilol treatment led to decreased troponin release at 4 months in breast cancer patients receiving ANT but did not impact LVEF reduction or prevent cardiovascular events.^23^ The antioxidant and pharmacological properties of ivabradine and its protection against the release of free radicals might contribute to its beneficial effects. The attenuation of the increase in troponin could improve the long-term prognosis of patients receiving high-dose ANT treatment.^24^

In 2006, Cardinale et al.^25^ reported ANT-induced LV dysfunction in patients receiving an average of 335 mg/m^2^ doxorubicin, and the LVEF decreased from 62.8% to 48.3% in the placebo group over 12 months. On the other hand, patients receiving enalapril during ANT maintained their LVEF and experienced no clinical complications. Those with elevated troponin levels throughout treatment had a more pronounced and persistent decrease in LVEF.

A few clinical trials have since evaluated the effectiveness of beta-blockers, neurohormonal antagonists, and atorvastatin in preventing ANT-related cardiovascular dysfunction.^23,26–37^ In most studies, neither a high prevalence of cardiovascular dysfunction nor benefits of pharmacological interventions were observed. Earlier trials with lower ANT doses, such as PRADA and CECCY, did not demonstrate protective effects of metoprolol and carvedilol against cardiotoxicity.^23,38^

The IPAC trial investigated the potential of ivabradine to mitigate GLS decline in patients receiving high doses of ANTs. Reduced GLS was common in patients after 6 months and persisted until the 12th month, although systolic dysfunction was rare. The hypothesis was that ivabradine would prevent ANT cardiotoxicity by reducing the HR and inducing antioxidant and antiapoptotic effects. However, ivabradine did not prevent cardiotoxicity, which was defined as a decline in GLS after 6 months. In a pilot study published in 2022 with patients with preserved LVEF and HR ≥75 bpm, ivabradine did not alter the diastolic function or BNP levels but significantly improved the GLS.^39^

The association between GLS and HR has been previously reported. Peverill et al.^40^ showed that GLS in patients with preserved LVEF was independently and inversely related to HR, and Kraigher-Krainer et al.^41^ demonstrated that worse GLS values were directly associated with higher HRs in patients with preserved LVEF. This informed the study’s primary endpoint choice on the basis of the mechanism of action of ivabradine.

Over 12 months of follow-up, patients who received ANT had a significant reduction in HR, but ivabradine did not increase this reduction in HR. This finding may have been due to the fixed dose of 5 mg BID in this study, without any titration to higher doses. Another possibility is that inhibitors and inducers of CYP3A4 may interact with ivabradine hydrochloride in cancer patients, potentially influencing the metabolism and pharmacokinetics of this drug to a clinically significant extent. We ensured reliable adherence in our study, as evidenced by the meticulous pill counting conducted at each visit. Thus, in this trial, we did not observe the expected reduction in HR with ivabradine, which might explain the neutral effects of the drug on anthracycline cardiotoxicity.

In the BEAUTIFUL study, ivabradine did not reduce death or hospitalization in patients with stable coronary artery disease and an LVEF <40%. However, a subgroup analysis revealed reduced hospitalizations for myocardial infarction and decreased need for revascularization in patients with a baseline HR of <70 bpm.^42^ The SHIFT study with 6558 HF patients with an LVEF <35% revealed that ivabradine, titrated up to 7.5 mg twice daily, reduced the HR, leading to fewer hospitalizations and HF-related deaths.^43^ In both studies, ivabradine was effective in patients with established systolic dysfunction, indicating the benefits of HR control in such patients. The ineffectiveness of ivabradine in our study might have stemmed from its preventive rather than therapeutic use.

In this study, ivabradine was well tolerated but did not effectively prevent GLS decline, a decreased LV ejection fraction, or clinical complications in ANT-treated patients, likely because of insufficient HR reduction. In 2023, Neilan et al.^33^ published the STOP-CA trial, which evaluated the efficacy of atorvastatin in preventing a decrease in LVEF in lymphoma patients. Over 12 months, a >10% decrease in LVEF to <55% was detected in 22% of the placebo patients, whereas a 9% decrease in LVEF was detected in the atorvastatin-treated group. The protective effect of ivabradine in vitro may be related to its anti-inflammatory, antioxidative, and antiapoptotic mechanisms. In the IPAC trial, while there was no significant reduction in the LVEF or decrease in the GLS, lower troponin release suggested that ivabradine may reduce myocardial injury (as indicated by troponin levels) but might not effectively prevent left ventricular systolic dysfunction in cancer patients.

Our investigation has several limitations. First, owing to its single-center design, it may not represent all patient demographics, despite being the first study to test ivabradine for cardiotoxicity. Second, the fixed ivabradine dose of 10 mg/day was not titrated, resulting in a failure to reduce the patient HR. This limitation could have hindered the ability to assess the effectiveness of ivabradine accurately in this study. Third, we conducted a 12-month follow-up analysis of patients to assess clinical outcomes, enabling the detection of differences between groups in later stages of the follow-up period. Fourth, although our study’s follow-up period was only 12 months, the possibility of observing long-term differences between groups in future analyses cannot be ruled out. Finally, one of the primary limitations of this trial pertains to its power. The sample size was calculated based on an anticipated incidence of cardiotoxicity of 40% with the use of ANT and an optimistic expectation of reducing this incidence to 20% with the introduction of ivabradine. While this expectation underpinned the trial design, it inherently limited the ability to detect more modest effects of ivabradine in this study.

In conclusion, ivabradine at a fixed 10 mg/day dose, did not have a definitive protective effect against cardiotoxicity in cancer patients receiving ANT therapy as initially hypothesized. However, intriguing observations regarding the potential of ivabradine to reduce myocardial injury by reducing troponin levels might be explored in future trials.

## Non-standard abbreviations and acronyms

ANT: anthracycline
CTRCD: cancer therapy-related cardiac dysfunction
EKG: electrocardiogram
HF: heart failure
HR: heart rate
LV: left ventricle
LVEF: left ventricular ejection fraction
NT-proBNP: N-terminal pro B-type natriuretic peptide
TTE: transthoracic echocardiogram
GLS: global longitudinal strain

## Acknowledgments

none.

## Data Availability Statement

The data that support the findings of this study are available from the corresponding author upon reasonable request.

